# The effectiveness and safety of rhythm control for atrial fibrillation in patients with end-stage or chronic kidney disease

**DOI:** 10.1101/2023.07.26.23293221

**Authors:** Dong-Seon Kang, Daehoon Kim, Eunsun Jang, Hee Tae Yu, Tae-Hoon Kim, Jae-Sun Uhm, Jung-Hoon Sung, Hui-Nam Pak, Moon-Hyoung Lee, Pil-Sung Yang, Boyoung Joung

## Abstract

**Background:** Although early rhythm control improves cardiovascular outcomes in patients with atrial fibrillation (AF), its use in patients with end-stage renal disease (ESRD) remains challenging. This study aimed to investigate the effectiveness and safety of rhythm control in patients with renal failure, including ESRD.

**Methods:** This population-based cohort study included 31,687 patients with AF who underwent rhythm or rate control between 2005 and 2015. Patients were categorized into three groups based on the estimated glomerular filtration rate: ESRD (<15ml/min/1.73m² or undergoing dialysis), 15– 60ml/min/1.73m², and ≥60ml/min/1.73m². The primary outcome consisted of cardiovascular death, ischemic stroke, heart failure-related hospitalization, and acute myocardial infarction.

**Results:** Among study population, 20,629 (65.1%) were male patients, with a median age of 63 years and a median follow-up period of 3.6 years. In the ESRD group, the comparative effectiveness of rhythm control was not significant (hazard ratio [HR] 0.97, 95% confidence interval [CI] 0.81 to 1.17). However, in the 15–60ml/min/1.73m² group, rhythm control was associated with a lower risk of the primary outcome than rate control (HR 0.85, 95% CI 0.74 to 0.98). This beneficial trend was consistently observed in the ≥60ml/min/1.73m² group (HR 0.87, 95% CI 0.80 to 0.93). No significant interaction was observed between renal function and treatment (p for interaction = 0.172). Rhythm control tended to have a significantly higher risk for the composite safety outcome than rate control in the ESRD group, with a significant renal function-by-treatment interaction (HR 1.29, 95% CI 1.11 to 1.50; p for interaction = 0.016).

**Conclusion:** In patients with renal failure, rhythm control was associated with better cardiovascular outcomes than rate control. However, the comparative effectiveness of rhythm control was less prominent in patients with ESRD, and the risk of adverse outcomes was higher than that of rate control. Therefore, rhythm control should be considered selectively in patients with renal failure.

**Clinical Perspective:** **What is new?**

- Among patients with relatively preserved renal function, rhythm control strategies were associated with a lower risk of primary outcome than rate control strategies. However, this beneficial trend was less prominent in patients with end-stage renal disease (ESRD).
- In addition, in patients with ESRD, unlike in patients with relatively preserved renal function, rhythm control strategies were significantly associated with a higher risk of composite safety outcome than rate control strategies.

**What are the clinical implications?**

- Recent major clinical trials have reported that early application of antiarrhythmic agents or catheter ablation in patients with atrial fibrillation can improve cardiovascular outcomes. However, patients with advanced renal failure, including ESRD, were largely excluded from these studies. As a result, the comparative effectiveness of rhythm control strategies could not be generalized to this specific AF population.
- This nationwide population-based study will assist in identifying appropriate patient selection based on renal function to ensure the benefits of rhythm control strategies.

## Introduction

Atrial fibrillation (AF) is a common form of arrhythmia, and its associated symptoms and stroke risk can be significantly improved through rate control strategies and anticoagulation therapy.^1, 2^ Recent studies have demonstrated that implementing rhythm control strategies within 1 year of AF diagnosis as part of an integrated management pathway is associated with a lower cardiovascular outcome risk.^3–7^

However, uniform implementation of rhythm control strategies as a first-line treatment for AF remains challenging. Proarrhythmic potential poses a significant hurdle in using antiarrhythmic drugs (AAD), and many of these drugs undergo renal metabolism or excretion. Consequently, current guidelines discourage the use of most AAD in patients with renal failure, particularly those with end-stage renal disease (ESRD).^8, 9^ Similarly, catheter ablation, which has been increasingly used and is known to have a potent antiarrhythmic effect than AAD, has been associated with reported major bleeding and cardiac tamponade in 5.4% and 3.2% of patients with ESRD, respectively.^9–11^ Indeed, concerns about the safety of rhythm control strategies have hindered physicians from properly utilizing them in real-world settings for patients with advanced renal failure, including ESRD.^12^ However, the exclusion of patients with an estimated glomerular filtration rate (eGFR) < 15 ml/min/1.73 m² or those undergoing dialysis from the most pivotal clinical trials assessing rhythm control strategies in AF has resulted in uncertain cardiovascular outcomes associated with these strategies in the specific AF population.^13–15^ With an aging population and increasing obesity rates, the prevalence of AF and renal failure is increasing, with reports indicating that 10–20% of patients with ESRD have AF.^9, 16^ Therefore, it is crucial to investigate the effectiveness and safety of rhythm control strategies in patients with renal failure, especially ESRD, to optimize their management. This study aimed to examine the influence of rhythm control strategies on cardiovascular outcomes in patients with AF across different renal function levels, including ESRD, by assessing the benefits and risks of rhythm control strategies to help select suitable patients.

## Methods

### Data sources

Data for this retrospective analysis were obtained from the National Health Insurance Service (NHIS) database in Korea. The NHIS is managed by the Korean government and covers the majority (97.1%) of the Korean population, with the remaining 3% receiving medical aid. The database includes information on sociodemographic factors, inpatient and outpatient services, prescriptions, and mortality and is accessible to the public on the NHIS website (http://nhiss.nhis.or.kr). This study was approved by the Institutional Review Board of the Yonsei University Health System (4-2022-1241). Owing to strict confidentiality guidelines, personal identification information was removed after cohort generation, and the requirement for informed consent was waived.

### Cohort design and study population

Between January 2005 and December 2015, we identified patients aged ≥ 18 years who were diagnosed with AF and received treatment using either rhythm or rate control strategies. eGFR was determined using the Chronic Kidney Disease Epidemiology Collaboration (CKD-EPI) equation.^17^ Among them, patients who did not have eGFR measured at baseline, whose disease duration at treatment initiation was more than 1 year after AF diagnosis, or who died within 30 days after the initial prescription or procedure were excluded from the study (Figure 1). The patients were classified into three groups based on the following eGFR ranges: ESRD (eGFR < 15 ml/min/1.73 m² or undergoing dialysis), 15–60 ml/min/1.73 m^2^, and ≥ 60 ml/min/1.73 m^2^. AF was diagnosed using the International Classification of Diseases, Tenth Revision (ICD-10) code I48. The accuracy of this diagnosis was previously confirmed in the NHIS database, with a positive predictive value of 94.1%.^18^ To investigate the use of rhythm or rate control strategies, a “new use” and “intention to treat” design was employed, as previously reported.^4, 19^ “New use” was defined as the absence of prior treatment records in the NHIS database. This database provides comprehensive information on drug prescriptions for the entire Korean population from January 1, 2002, enabling a retrospective verification period of at least 9.5 years.

**Figure 1.**
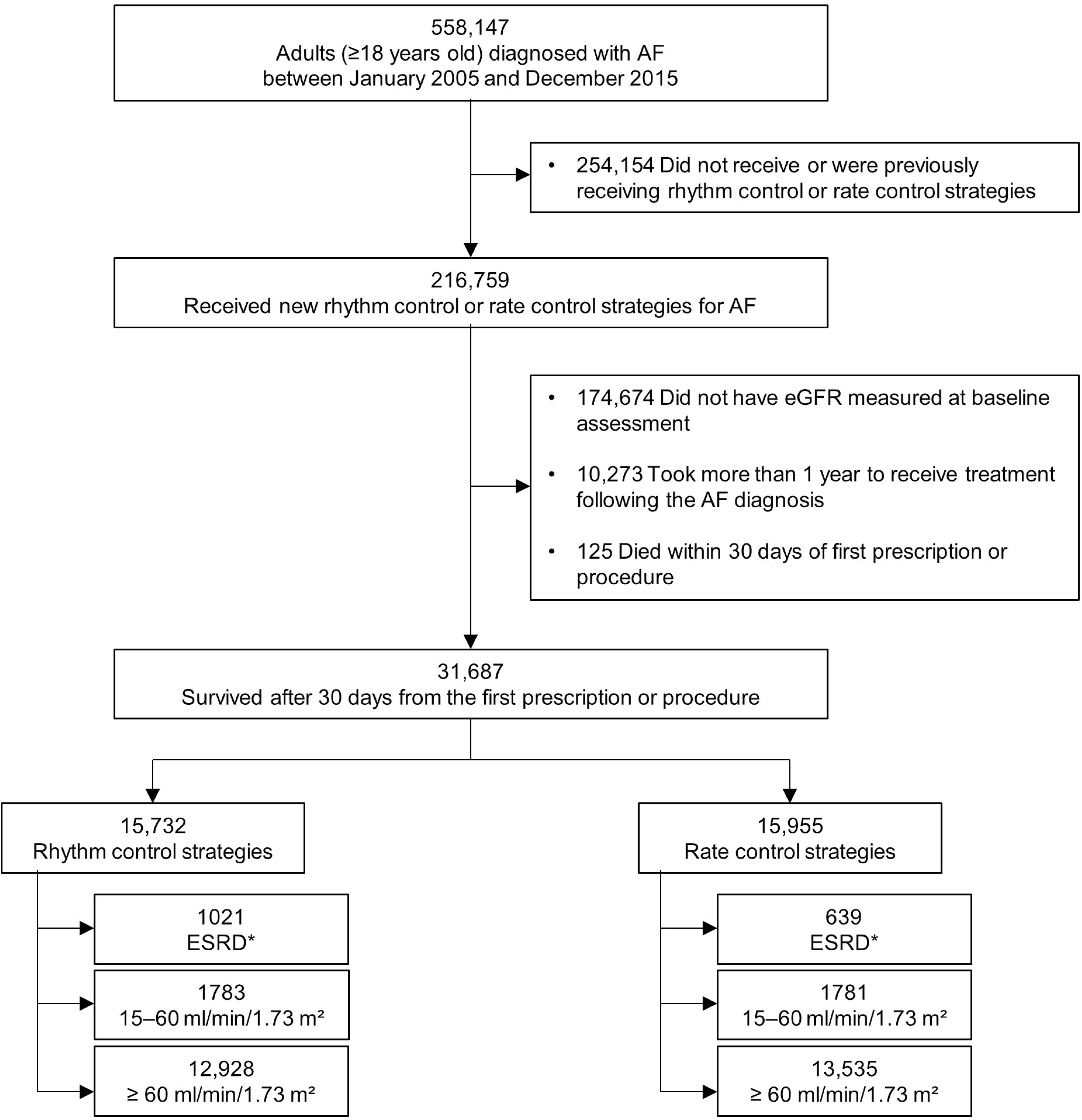
Flow chart of enrollment and analysis of the study population. *ESRD was defined as an estimated glomerular filtration rate less than 15 ml/min/1.73 m^2^ or undergoing dialysis. AF, atrial fibrillation; eGFR, estimated glomerular filtration rate; ESRD, end-stage renal disease.

We defined “intention to treat” with rhythm control strategies as the prescription of any AAD for more than 20 days or the performance of an ablation procedure for AF within 30 days of the initial prescription. “Intention to treat” with rate control strategies was defined as the prescription of any rate control drug for more than 20 days within 30 days of the initial prescription, without any prescriptions of AAD or ablation procedures during this period. The drugs used for rhythm and rate control strategies, as well as the claim codes for ablation procedures, are presented in Supplementary Table 1.

### Outcome and covariates

The primary outcome of this study was a composite of cardiovascular death, ischemic stroke, heart failure-related hospitalization, or acute myocardial infarction. We also assessed each component of the primary outcome individually during the follow-up period, using the same endpoints as in the EAST-AFNET 4 trial. Additionally, we defined a composite safety outcome that consisted of all cause death, intracranial hemorrhage, gastrointestinal bleeding, and serious adverse event related to rhythm control. Supplementary Table 2 presents detailed definitions of the outcomes. Our analysis did not include recurrent events. The follow-up of the study outcomes was initiated 30 days after the initial prescription or procedure and continued until the end of the follow-up period (December 31, 2016) or until the patient’s death. Baseline comorbidity information was collected using inpatient and outpatient hospital diagnoses and pharmacy claims within the look-back period starting from January 1, 2002 (Supplementary Table 1). Patients’ baseline relative economic status was determined based on their health insurance premiums in the index year. Concurrent medication use was confirmed through claims in the NHIS database and was defined as a prescription for more than a 20-day supply within 30 days of initiating rhythm or rate control strategies. None of the variables had any missing data.

### Statistical analysis

The baseline characteristics of the patients were described using descriptive statistics. To address the differences in the baseline characteristics between the study treatment strategies, we employed propensity overlap weighting. The model included several variables, such as sociodemographic factors, time since AF diagnosis, level of care for the initial prescription, clinical risk scores, comorbidities, and concurrent medication use. Supplementary Figure 1 shows the distribution of the propensity scores before and after overlap weighting in each renal function group. Balance was evaluated using standardized differences, with a threshold of 0.1 indicating an imbalance. Weighted incidence rates were calculated by dividing the weighted number of clinical events during the follow-up period by 100 person-years at risk. To estimate the relative hazards of the outcomes of interest, we employed a Fine and Gray competing risk regression model with an interaction term for the renal function group and treatment strategy (rhythm or rate control strategies), accounting for all cause death as a competing event. Covariates that remained unbalanced after weighting were included as cofactors in a competing risk regression analysis. The proportional hazard assumption was evaluated based on Schoenfeld residuals, and no violations were detected. To investigate the eGFR-dependent effect of rhythm control strategies on the primary outcome, we employed a Cox proportional hazards model on the weighted study participants, incorporating an interaction term for eGFR (assessed as a continuous variable and modeled as a cubic spline) and treatment strategy. Statistical significance was defined as a two-sided P < 0.05. All statistical analyses were conducted using R version 4.2.1 (The R Foundation, www.R-project.org).

### Sensitivity analyses

First, subgroup analyses were performed on the primary outcome, including sex, age, baseline renal function, and the Charlson comorbidity index. Interaction tests were conducted for all the subgroups. Second, all variables used in the propensity score calculation were included as covariates in the competing risk regression models to account for residual imbalance after overlap weighting. Third, we used one-to-one propensity matching without replacement with a caliper of 0.01 to adjust for the differences in baseline characteristics between the two AF treatment strategies. Fourth, we conducted a falsification analysis to assess systematic bias in our study by measuring the effects of AF treatment strategies on 22 prespecified endpoints with true hazard ratios (HR) of 1. Detailed definitions of the falsification endpoints are presented in Supplementary Table 3.

## Results

### Baseline characteristics

After exclusion, the study population consisted of 31,687 patients with a median age of 63 (interquartile range 54–72) years. Among them, 1660 (5.2%) constituted the ESRD group, 3564 (11.2%) exhibited an eGFR ranging from 15–60 ml/min/1.73 m², and the remaining 26,463 (83.5%) exhibited an eGFR of ≥ 60 ml/min/1.73 m². The detailed baseline characteristics based on renal function with eGFR and AF treatment strategies are presented in Table 1. Compared with rate control strategies, rhythm control strategies were associated with higher incomes and higher rates of comorbidities, including hypertension, diabetes mellitus, dyslipidemia, ischemic stroke, myocardial infarction, and peripheral arterial disease across renal function. Moreover, as renal function declined, the burden of comorbidities increased. After weighting, all variables were balanced between the two AF treatment strategies (Supplementary Table 4). Among rhythm control strategies, amiodarone and propafenone were the most frequently used drugs, followed by flecainide and pilsicainide (Figure 2). Ablation was initially employed as the initial strategy in 6 (0.6%), 5 (0.3%), and 75 (0.6%) patients among the ESRD, 15–60 ml/min/1.73 m², and ≥ 60 ml/min/1.73 m² groups, respectively. However, it was performed during the follow-up period in 38 (2.3%), 86 (4.8%), and 1349 (10.4%) patients, respectively.

**Figure 2.**
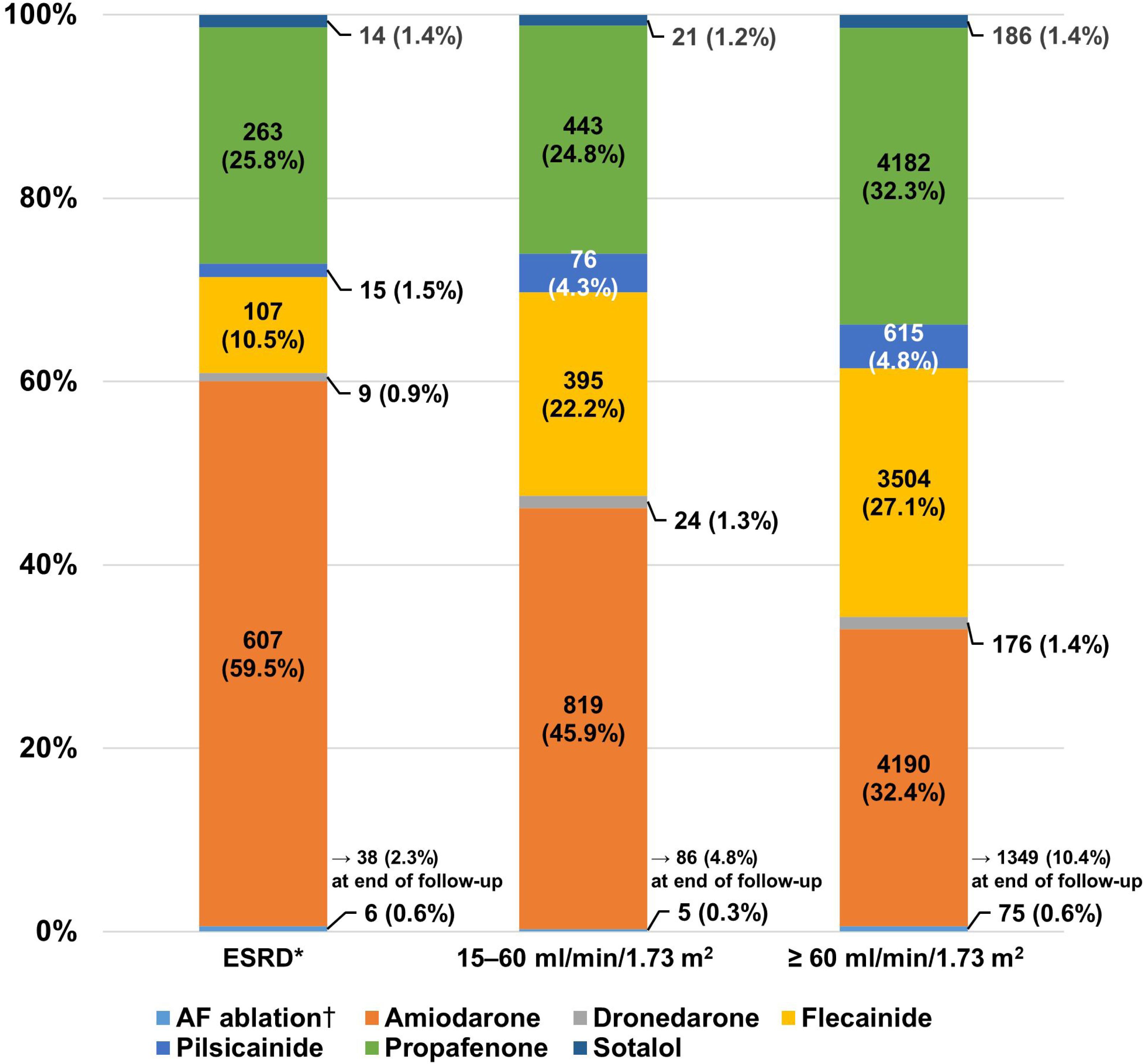
Initial choice in patients who underwent rhythm control strategies according to renal function. * ESRD was defined as an estimated glomerular filtration rate less than 15 ml/min/1.73 m^2^ or undergoing dialysis. †If the ablation procedure was performed within 30 days of initiating the rhythm control strategy, it was considered as an initial choice. AF, atrial fibrillation; ESRD, end-stage renal disease.

**Table 1.** Baseline characteristics before overlap weighting according to renal function.

|  | ESRD* |  | 15–60 ml/min/1.73 m<sup>2</sup> |  | ≥ 60 ml/min/1.73 m<sup>2</sup> |  |
| --- | --- | --- | --- | --- | --- | --- |
|  | Rhythm control strategies<br>(N=1021) | Rate control strategies<br>(N=639) | Rhythm control strategies<br>(N=1783) | Rate control strategies<br>(N=1781) | Rhythm control strategies<br>(N=12,928) | Rate control strategies<br>(N=13,535) |
| Age, years | 63 (55–71) | 60 (49–68) | 72 (66–77) | 74 (67–79) | 61 (53–69) | 62 (54–71) |
| <65 years | 55.7 | 63.4 | 21.6 | 18.7 | 61.1 | 56.0 |
| 65–74 years | 30.4 | 27.2 | 38.8 | 35.8 | 28.4 | 28.7 |
| ≥75 years | 13.9 | 9.4 | 39.5 | 45.5 | 10.5 | 15.3 |
| Male | 61.7 | 62.0 | 53.9 | 59.2 | 67.7 | 65.3 |
| Baseline eGFR (mL/min/1.73 m<sup>2</sup>) | 6.0 (4.6–9.9) | 6.0 (4.7–8.9) | 53.2 (47.1–56.9) | 53.8 (47.8–57.0) | 83.8 (72.6–94.2) | 84.4 (73.1–95.0) |
| AF duration (months) | 0.0 (0.0–0.8) | 0.0 (0.0–1.4) | 0.0 (0.0–0.8) | 0.0 (0.0–0.2) | 0.0 (0.0–0.7) | 0.0 (0.0–0.1) |
| High tertile of income | 39.0 | 34.7 | 51.1 | 44.1 | 47.7 | 41.3 |
| No of OPD visits ≥ 12/years | 86.9 | 88.6 | 86.8 | 81.5 | 79.3 | 76.0 |
| Living in metropolitan areas | 47.0 | 47.1 | 42.5 | 38.6 | 43.6 | 38.9 |
| Level of care initiating treatment |  |  |  |  |  |  |
| Primary | 6.0 | 10.0 | 9.0 | 17.7 | 9.4 | 19.3 |
| Secondary | 40.9 | 44.9 | 45.2 | 49.9 | 42.5 | 50.2 |
| Tertiary | 53.1 | 45.1 | 45.8 | 32.4 | 48.0 | 30.5 |
| Risk scores |  |  |  |  |  |  |
| CHA<sub>2</sub>DS<sub>2</sub>-VASc score | 4 (3–6) | 3 (2–4) | 4 (3–6) | 4 (2–5) | 2 (1–4) | 2 (1–3) |
| mHAS-BLED score† | 3 (3–4) | 3 (2–4) | 3 (2–4) | 3 (2–3) | 2 (1–3) | 2 (1–3) |
| Charlson comorbidity index | 8 (5–10) | 6 (3–8) | 5 (3–7) | 4 (2–6) | 3 (2–5) | 3 (1–4) |
| Hospital Frailty Risk score | 7.5 (3.7–13.1) | 5.0 (1.4–9.4) | 2.5 (0.0–6.7) | 1.8 (0.0–5.7) | 0.9 (0.0–3.5) | 0.8 (0.0–3.6) |
| Medical history |  |  |  |  |  |  |
| Hypertension | 91.1 | 76.7 | 82.5 | 64.2 | 57.9 | 45.3 |
| Diabetes mellitus | 57.2 | 48.2 | 29.6 | 23.4 | 16.7 | 14.5 |
| Dyslipidemia | 85.8 | 69.2 | 84.2 | 67.4 | 72.5 | 62.0 |
| Heart failure | 51.9 | 37.7 | 44.7 | 40.8 | 27.7 | 28.7 |
| Ischemic stroke | 27.4 | 18.2 | 27.6 | 22.7 | 13.3 | 12.2 |
| Transient ischemic attack | 9.9 | 4.7 | 11.8 | 9.4 | 7.0 | 5.2 |
| Intracranial hemorrhage | 4.1 | 3.1 | 2.8 | 1.6 | 1.4 | 1.8 |
| Myocardial infarction | 22.4 | 15.3 | 12.2 | 7.9 | 5.7 | 4.7 |
| Peripheral arterial disease | 24.7 | 14.2 | 20.1 | 13.0 | 11.3 | 7.8 |
| Valvular heart disease | 7.0 | 5.2 | 4.8 | 4.7 | 5.1 | 4.6 |
| Hyperthyroidism | 15.2 | 9.2 | 10.3 | 8.0 | 11.4 | 10.7 |
| Hypothyroidism | 18.5 | 11.7 | 13.7 | 8.3 | 10.3 | 8.1 |
| Malignancy | 25.5 | 19.2 | 26.9 | 25.2 | 20.7 | 19.8 |
| COPD | 31.0 | 22.5 | 38.0 | 34.5 | 23.4 | 23.0 |
| Chronic liver disease | 50.8 | 42.3 | 45.4 | 37.8 | 42.0 | 38.5 |
| Hypertrophic cardiomyopathy | 0.7 | 0.2 | 1.8 | 0.5 | 1.0 | 0.4 |
| Osteoporosis | 39.7 | 26.1 | 42.2 | 34.6 | 23.2 | 22.9 |
| Sleep apnea | 0.5 | 0.2 | 0.4 | 0.4 | 0.8 | 0.4 |
| Concurrent medication‡ |  |  |  |  |  |  |
| Oral anticoagulant | 26.3 | 12.4 | 43.1 | 30.9 | 38.7 | 22.9 |
| Warfarin | 25.9 | 11.7 | 36.6 | 27.0 | 32.7 | 20.5 |
| Direct oral anticoagulant | 0.6 | 0.8 | 8.5 | 4.6 | 7.4 | 3.1 |
| Beta blocker | 31.1 | 35.4 | 30.2 | 42.9 | 29.3 | 48.5 |
| Non-DHP CCB | 6.2 | 6.7 | 6.5 | 11.2 | 8.1 | 10.4 |
| Aspirin | 34.6 | 24.4 | 30.3 | 36.7 | 33.4 | 35.4 |
| P<sub>2</sub>Y<sub>12</sub> inhibitor | 15.7 | 10.0 | 11.4 | 11.8 | 8.1 | 8.4 |
| Statin | 21.3 | 15.8 | 23.7 | 19.7 | 18.8 | 17.5 |
| DHP CCB | 23.1 | 22.7 | 14.8 | 9.4 | 9.2 | 6.8 |
| ACEi/ARB | 32.2 | 26.9 | 33.4 | 33.9 | 24.3 | 26.1 |
| Loop/thiazide diuretics | 17.7 | 22.8 | 28.9 | 33.2 | 16.5 | 23.9 |
| K<sup>+</sup> sparing diuretics | 1.6 | 2.8 | 8.2 | 11.5 | 5.2 | 8.9 |
| Alpha blocker | 3.1 | 5.3 | 2.4 | 1.9 | 1.1 | 1.3 |

Values are presented as median (interquartile range) or %.
\*ESRD was defined as an estimated glomerular filtration rate less than 15 ml/min/1.73 m<sup>2</sup> or undergoing dialysis.
†Modified HAS-BLED=hypertension, 1 point; age >65 years, 1 point; history of stroke, 1 point; history of bleeding or predisposition, 1 point; liable international normalized ratio, not assessed; ethanol or drug abuse, 1 point; drug predisposing to bleeding, 1 point.
‡Concurrent medication use was defined as a prescription for more than a 20-day supply within 30 days of initiating rhythm or rate control strategies.
ACEi, angiotensin converting enzyme inhibitor; AF, atrial fibrillation; ARB, angiotensin receptor blocker; CCB, calcium channel blockers; COPD, chronic obstructive pulmonary disease; DHP, dihydropyridine; eGFR, estimated glomerular filtration rate; ESRD, end-stage renal disease; OPD, outpatient department.

### Comparative effectiveness and safety of rhythm control strategies in patients with renal failure

During a median follow-up period of 3.6 (interquartile range 2.1–5.4) years, the comparative effectiveness of rhythm control strategies was less prominent in the ESRD group (weighted incidence rate of 9.94 events per 100 person-years in rhythm control strategies vs. 9.42 events per 100 person-years in rate control strategies; HR 0.97, 95% confidence interval [CI] 0.81 to 1.17) (Figures 3 and 4). However, in the 15–60 ml/min/1.73 m^2^ group, rhythm control strategies were associated with a lower risk of the primary outcome than rate control strategies (weighted incidence rate of 7.69 events per 100 person-years in rhythm control strategies vs. 9.37 events per 100 person-years in rate control strategies; HR 0.85, 95% CI 0.74 to 0.98). This beneficial trend was consistently observed in the ≥ 60 ml/min/1.73 m^2^ group (weighted incidence rate of 3.64 events per 100 person-years in rhythm control strategies vs. 4.21 events per 100 person-years in rate control strategies; HR 0.87, 95% CI 0.80 to 0.93). No significant interaction was identified between the renal function group and the effect of rhythm control strategies on the primary outcome (*p* for interaction = 0.172).

**Figure 3.**
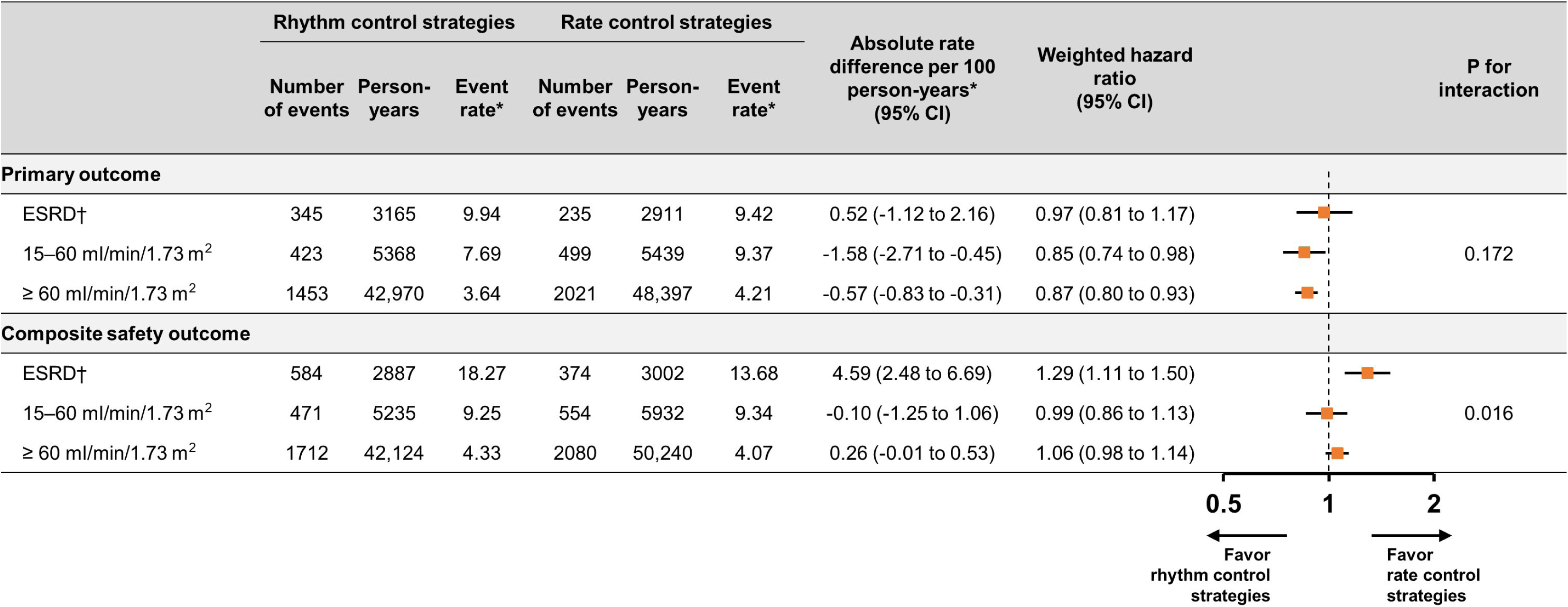
The primary outcome and composite safety outcome in patients who received rhythm and rate control strategies. *Weighted incidence rate (per 100 person-years) after overlap weighting was applied. †ESRD was defined as an estimated glomerular filtration rate less than 15 ml/min/1.73 m^2^ or undergoing dialysis. CI, confidence interval; ESRD, end-stage renal disease.

**Figure 4.**
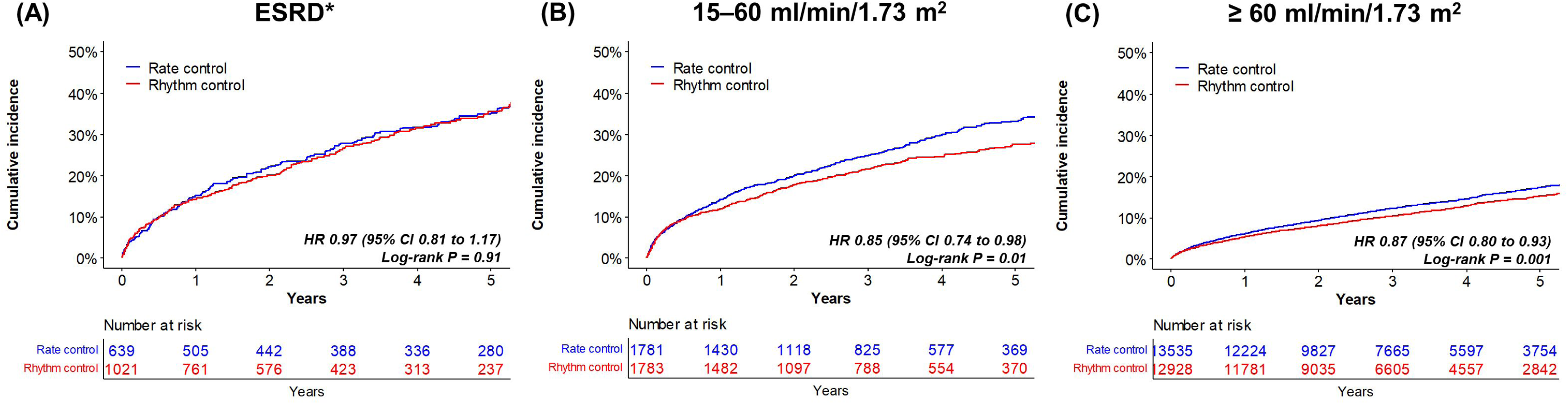
Weighted cumulative incidence curves for the primary outcome according to renal function. The results are presented for three renal function groups: ESRD (A), 15–60 ml/min/1.73 m² (B), and ≥ 60 ml/min/1.73 m² (C). *ESRD was defined as an estimated glomerular filtration rate less than 15 ml/min/1.73 m^2^ or undergoing dialysis. I, confidence interval; ESRD, end-stage renal disease; HR, hazard ratio.

The results of the individual components of the primary outcome are presented in Table 2 and Supplementary Figure 2. These findings demonstrated that the beneficial trend of rhythm control strategies primarily originated from the effects on ischemic stroke across all renal function groups (HR 0.72, 95% CI 0.53 to 0.98 in the ESRD group; HR 0.65, 95% CI 0.53 to 0.81 in the 15–60 ml/min/1.73 m^2^ group; HR 0.88, 95% CI 0.79 to 0.98 in the ≥ 60 ml/min/1.73 m^2^ group). Conversely, the significantly higher risk of cardiovascular death in the ESRD group contributed to the diminished comparative effectiveness of rhythm control strategies (weighted incidence rate of 4.36 events per 100 person-years in rhythm control strategies vs. 2.57 events per 100 person-years in rate control strategies; HR 1.58, 95% CI 1.15 to 2.15).

**Table 2.** Individual components of the primary outcome in patients who received rhythm and rate control strategies.

|  | Number of<br>events | Person-years | Event<br>rate* | Number<br>of events | Person-years | Event<br>rate* | Absolute rate<br>difference per 100<br>person-years*<br>(95% CI) | Weighted hazard<br>ratio<br>(95% CI) |
| --- | --- | --- | --- | --- | --- | --- | --- | --- |
| **ESRD†** | **Rhythm control strategies**<br>**(N=1021)** |  |  | **Rate control strategies**<br>**(N=639)** |  |  |  |  |
| Cardiovascular death | 175 | 3629 | 4.36 | 78 | 3479 | 2.57 | 1.78 (0.90 to 2.67) | 1.58 (1.15 to 2.15) |
| Ischemic stroke | 110 | 3438 | 2.94 | 100 | 3213 | 3.61 | -0.67 (-1.59 to 0.25) | 0.72 (0.53 to 0.98) |
| Heart failure-related hospitalization | 115 | 3390 | 3.25 | 101 | 3184 | 3.95 | -0.71 (-1.68 to 0.26) | 0.72 (0.53 to 0.98) |
| Acute myocardial infarction | 57 | 3530 | 1.45 | 48 | 3362 | 1.60 | -0.15 (-0.76 to 0.46) | 0.81 (0.51 to 1.27) |
| **15–60 ml/min/1.73 m<sup>2</sup>** | **Rhythm control strategies**<br>**(N=1783)** |  |  | **Rate control strategies**<br>**(N=1781)** |  |  |  |  |
| Cardiovascular death | 141 | 6039 | 2.44 | 169 | 6329 | 2.62 | -0.18 (-0.75 to 0.39) | 0.95 (0.74 to 1.21) |
| Ischemic stroke | 172 | 5709 | 2.83 | 245 | 5839 | 4.36 | -1.53 (-2.24 to -0.83) | 0.65 (0.53 to 0.81) |
| Heart failure-related hospitalization | 188 | 5683 | 3.26 | 216 | 5881 | 3.62 | -0.36 (-1.05 to 0.33) | 0.92 (0.74 to 1.14) |
| Acute myocardial infarction | 29 | 5975 | 0.39 | 40 | 6248 | 0.65 | -0.26 (-0.53 to -0.01) | 0.60 (0.36 to 0.99) |
| **≥ 60 ml/min/1.73 m<sup>2</sup>** | **Rhythm control strategies**<br>**(N=12,928)** |  |  | **Rate control strategies**<br>**(N=13,535)** |  |  |  |  |
| Cardiovascular death | 331 | 45,656 | 0.82 | 473 | 52,478 | 0.84 | -0.02 (-0.14 to 0.10) | 0.98 (0.84 to 1.14) |
| Ischemic stroke | 702 | 44,234 | 1.71 | 961 | 50,428 | 1.94 | -0.23 (-0.41 to -0.06) | 0.88 (0.79 to 0.98) |
| Heart failure-related hospitalization | 595 | 44,397 | 1.48 | 904 | 50,418 | 1.80 | -0.32 (-0.48 to -0.15) | 0.82 (0.73 to 0.93) |
| Acute myocardial infarction | 100 | 45,459 | 0.22 | 118 | 52,241 | 0.24 | -0.01 (-0.08 to 0.05) | 0.94 (0.70 to 1.26) |

\*Weighted incidence rate (per 100 person-years) after overlap weighting was applied.
†ESRD was defined as an estimated glomerular filtration rate less than 15 ml/min/1.73 m<sup>2</sup> or undergoing dialysis.
CI, confidence interval; ESRD, end-stage renal disease.

The association between rhythm control strategies and the composite safety outcome varied, depending on the degree of renal function decline (Table 3 and Figure 3). Rhythm control strategies tended to have a higher risk of the composite safety outcome than rate control strategies in the ESRD group (weighted incidence rate of 18.27 events per 100 person-years in rhythm control strategies vs. 13.68 events per 100 person-years in rate control strategies; HR 1.29, 95% CI 1.11 to 1.50). However, no significant differences in the risk of the composite safety outcome were observed between rhythm and rate control strategies in the 15–60 ml/min/1.73 m^2^ (weighted incidence rate of 9.25 events per 100 person-years in rhythm control strategies vs. 9.34 events per 100 person-years in rate control strategies; HR 0.99, 95% CI 0.86 to 1.13) and the ≥ 60 ml/min/1.73 m^2^ groups (weighted incidence rate of 4.33 events per 100 person-years in rhythm control strategies vs. 4.07 events per 100 person-years in rate control strategies; HR 1.06, 95% CI 0.98 to 1.14). A significant interaction was observed between the renal function group and the effect of rhythm control strategies on the composite safety outcome (*p* for interaction = 0.016).

**Table 3.**
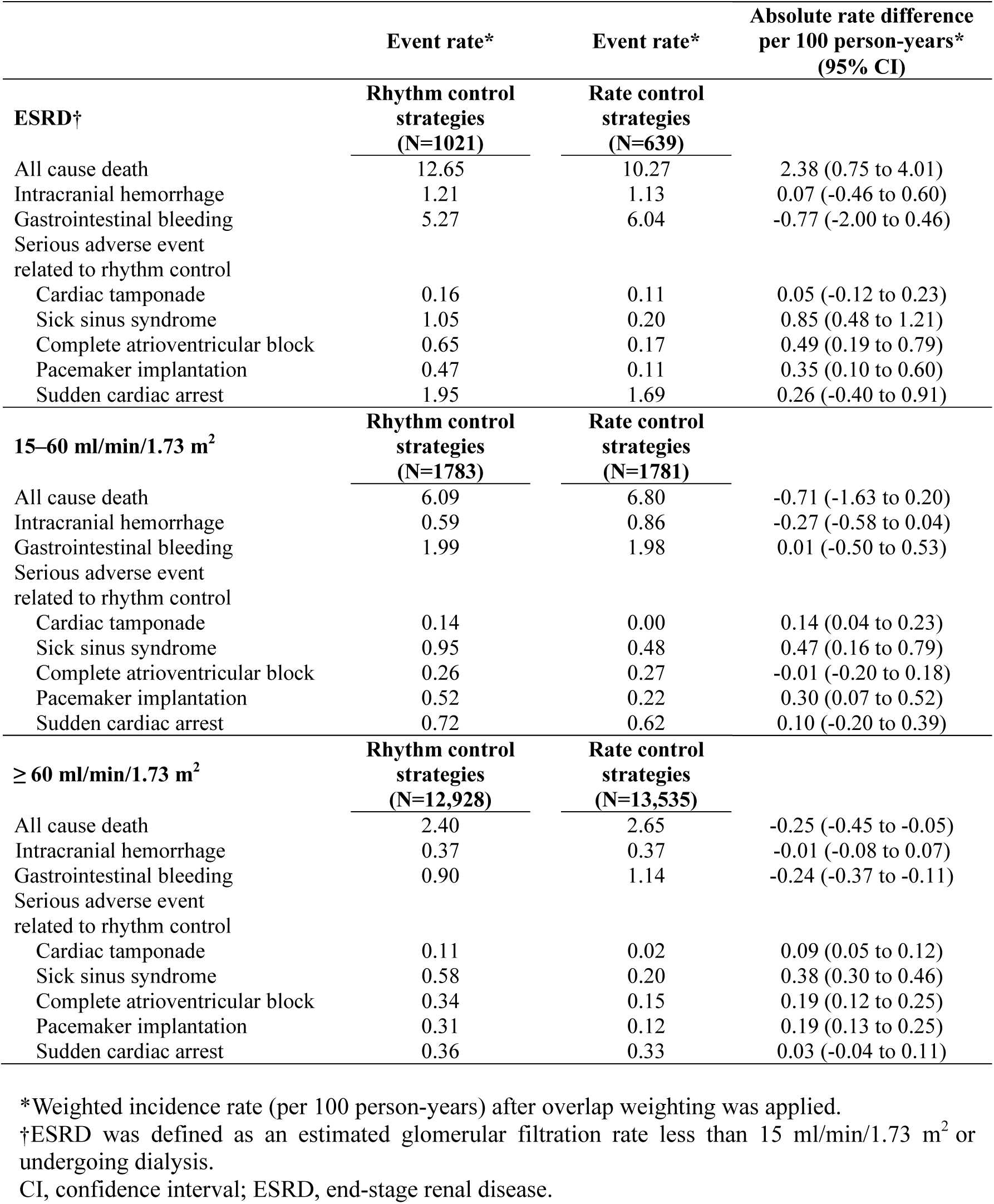
Individual components of the composite safety outcome (presented as rates per 100 person-years after overlap weighting) in patients who received rhythm and rate control strategies.

|  | Event rate* | Event rate* | Absolute rate difference<br>per 100 person-years*<br>(95% CI) |
| --- | --- | --- | --- |
|  | Rhythm control<br>strategies<br>(N=1021) | Rate control<br>strategies<br>(N=639) |  |
| **ESRD†** |  |  |  |
| All cause death | 12.65 | 10.27 | 2.38 (0.75 to 4.01) |
| Intracranial hemorrhage | 1.21 | 1.13 | 0.07 (-0.46 to 0.60) |
| Gastrointestinal bleeding | 5.27 | 6.04 | -0.77 (-2.00 to 0.46) |
| Serious adverse event<br>related to rhythm control |  |  |  |
| Cardiac tamponade | 0.16 | 0.11 | 0.05 (-0.12 to 0.23) |
| Sick sinus syndrome | 1.05 | 0.20 | 0.85 (0.48 to 1.21) |
| Complete atrioventricular block | 0.65 | 0.17 | 0.49 (0.19 to 0.79) |
| Pacemaker implantation | 0.47 | 0.11 | 0.35 (0.10 to 0.60) |
| Sudden cardiac arrest | 1.95 | 1.69 | 0.26 (-0.40 to 0.91) |
| **15–60 ml/min/1.73 m<sup>2</sup>** | **Rhythm control**<br>**strategies**<br>**(N=1783)** | **Rate control**<br>**strategies**<br>**(N=1781)** |  |
| All cause death | 6.09 | 6.80 | -0.71 (-1.63 to 0.20) |
| Intracranial hemorrhage | 0.59 | 0.86 | -0.27 (-0.58 to 0.04) |
| Gastrointestinal bleeding | 1.99 | 1.98 | 0.01 (-0.50 to 0.53) |
| Serious adverse event<br>related to rhythm control |  |  |  |
| Cardiac tamponade | 0.14 | 0.00 | 0.14 (0.04 to 0.23) |
| Sick sinus syndrome | 0.95 | 0.48 | 0.47 (0.16 to 0.79) |
| Complete atrioventricular block | 0.26 | 0.27 | -0.01 (-0.20 to 0.18) |
| Pacemaker implantation | 0.52 | 0.22 | 0.30 (0.07 to 0.52) |
| Sudden cardiac arrest | 0.72 | 0.62 | 0.10 (-0.20 to 0.39) |
| **≥ 60 ml/min/1.73 m<sup>2</sup>** | **Rhythm control**<br>**strategies**<br>**(N=12,928)** | **Rate control**<br>**strategies**<br>**(N=13,535)** |  |
| All cause death | 2.40 | 2.65 | -0.25 (-0.45 to -0.05) |
| Intracranial hemorrhage | 0.37 | 0.37 | -0.01 (-0.08 to 0.07) |
| Gastrointestinal bleeding | 0.90 | 1.14 | -0.24 (-0.37 to -0.11) |
| Serious adverse event<br>related to rhythm control |  |  |  |
| Cardiac tamponade | 0.11 | 0.02 | 0.09 (0.05 to 0.12) |
| Sick sinus syndrome | 0.58 | 0.20 | 0.38 (0.30 to 0.46) |
| Complete atrioventricular block | 0.34 | 0.15 | 0.19 (0.12 to 0.25) |
| Pacemaker implantation | 0.31 | 0.12 | 0.19 (0.13 to 0.25) |
| Sudden cardiac arrest | 0.36 | 0.33 | 0.03 (-0.04 to 0.11) |

\*Weighted incidence rate (per 100 person-years) after overlap weighting was applied.
†ESRD was defined as an estimated glomerular filtration rate less than 15 ml/min/1.73 m<sup>2</sup> or undergoing dialysis.
CI, confidence interval; ESRD, end-stage renal disease.

### Changes in comparative effectiveness of rhythm control strategies according to eGFR

The linear correlation between the comparative effectiveness of rhythm control strategies on the primary outcome and baseline eGFR, assessed as a continuous variable, is shown in Supplementary Figure 3. Rhythm control strategies demonstrated a significant benefit compared to rate control strategies, although this benefit gradually diminished with decreasing eGFR and became statistically insignificant when eGFR dropped below 30 ml/min/1.73 m².

### Sensitivity analyses

In the ESRD group, no significant interactions were observed between rhythm control strategies for the primary outcome and age, sex, baseline renal function, or the Charlson comorbidity index (Supplementary Figure 4). In the 15–60 ml/min/1.73 m^2^ group, the results were similar across all subgroups except for sex (p for interaction = 0.039) (Supplementary Figure 5). Comparative effectiveness of rhythm control strategies for the primary outcome was demonstrated in female patients (HR 0.72, 95% CI 0.59 to 0.89), but not in male patients (HR 0.89, 95% CI 0.81 to 1.19). Significant treatment-by-subgroup interactions were not demonstrated in the ≥ 60 ml/min/1.73 m^2^ group (Supplementary Figure 6).

The results from the competing risk regression, including all variables used in the propensity score calculation as covariates and one-to-one propensity matching, were consistent with the main findings (Supplementary Table 5). Regarding the analyses of the 22 falsification endpoints, the 95% CIs for the associations between rhythm control strategies and each endpoint included one for all endpoints (Supplementary Table 6).

## Discussion

In the present study, when rhythm control strategies were applied within 1 year of AF diagnosis in patients with renal failure, the risk of the primary composite outcome was significantly lower for rhythm control strategies than that for rate control strategies. This trend primarily originated from a reduced risk of ischemic stroke, which is consistent with the findings of previous major clinical trials.^3, 4, 15^ However, in patients with ESRD, the comparative effectiveness of rhythm control strategies was less prominent. Simultaneously, while the remaining two renal function groups demonstrated similar safety outcomes between the rhythm and rate control strategies, the ESRD group demonstrated a significantly higher risk associated with composite safety outcomes for rhythm control strategies compared to rate control strategies.

Patients with advanced renal failure, including ESRD, are underrepresented in randomized controlled trials.^3, 20^ The EAST-AFNET 4 trial, which demonstrated the effectiveness of rhythm control strategies within 1 year of AF diagnosis, excluded patients with ESRD and only included a modest number (≈13%) of patients with renal failure.^3^ In the subgroup analysis, no significant interaction was observed for the effectiveness of rhythm versus rate control strategies, even at eGFR ranging from 15–59 ml/min/1.73 m^2^. The CASTLE-AF trial, which demonstrated the association between catheter ablation and improved mortality in patients with concurrent AF and heart failure, also excluded patients with ESRD.^20^ Furthermore, the trial did not provide information regarding the inclusion of patients with eGFR < 60 ml/min/1.73 m² or whether a significant renal function-by-treatment interaction was observed in their study.

Recent studies have reported a high rate of adverse events related to rhythm control strategies in patients with renal failure. Bansal et al. reported that among patients with eGFR < 60 ml/min/1.73 m^2^, 27.6% experienced adverse events, with torsades de pointes occurring in 1.4% of cases.^21^ Furthermore, patients with eGFR < 15 ml/min/1.73 m^2^ exhibited a 43% higher risk of adverse events than those with eGFR > 60 ml/min/1.73 m^2^. Catheter ablation, another component of rhythm control strategies, has been reported to result in major complications, defined as requiring additional intervention or prolonged hospitalization within 3 months in 10% of patients with ESRD.^9^ By providing and comparing the benefits and risks of rhythm control strategies with those of rate control strategies, our study may assist in the meticulous application of rhythm control strategies in daily clinical practice.

In this study, the higher composite safety risk of rhythm control strategies in patients with ESRD primarily stemmed from an increased risk of cardiovascular death and bradyarrhythmias, including sick sinus syndrome. This result is supported by the increased proarrhythmic risk associated with impaired renal function because the metabolism of AAD, which accounted for most of the initial rhythm control strategies, largely depends on renal function.^8^ Moreover, the proarrhythmic potential is particularly prominent in the presence of structural heart disease.^22^ ESRD acts as a risk factor for significant coronary disease or left ventricular hypertrophy by promoting atherosclerosis and myocardial fibrosis through mechanisms, such as acid-base imbalance, altered calcium metabolism, and increased oxidative stress.^23–25^ Class IC AAD, which accounted for 37.8% of the rhythm control strategies in the ESRD group, is associated with unfavorable cardiovascular outcomes when prescribed in the presence of structural abnormalities.^26, 27^ In addition, myocardial electrical instability accompanies large and rapid shifts in fluid or electrolytes during hemodialysis.^28^ These factors can partially explain the poor cardiovascular outcomes in patients with ESRD who underwent rhythm control strategies.

Furthermore, during the period when patients were enrolled in the NHIS cohort, warfarin was the main prescribed anticoagulant. In addition, warfarin continues to be the first-line recommendation over direct anticoagulants in patients with ESRD.^29^ Considering that amiodarone, which potentiates the anticoagulant effect of warfarin, is the most common drug used in rhythm control strategy, the risk of composite safety outcomes, including intracranial hemorrhage and gastrointestinal bleeding, may have been overestimated, particularly in patients with ESRD.^30, 31^ This also indicates the need for a reevaluation of bleeding risk and the benefits and risks of rhythm control strategies in patients receiving AF treatments in the current era, when the role of direct anticoagulants and catheter ablation has become important.^32^

The present study has several limitations. First, owing to the use of a claims-based database, the burden of AF, its specific type, and accompanying symptoms could not be evaluated. Additionally, the presence of unmeasured confounding variables, including the quality of anticoagulation therapy or lifestyle factors, such as alcohol intake, was not considered. This prevented the establishment of causal relationships based on the findings of this observational study. However, the results of the falsification analysis demonstrated a low likelihood of significant systematic bias, and sufficient propensity score overlap was observed between the rhythm and rate control strategy groups. Second, the reliance on ICD-10 codes for diagnosing comorbidities and outcomes introduced the possibility of misdiagnosis owing to coding errors. To address this issue, we applied previously validated definitions from previous studies utilizing the Korean NHIS database.^18, 33^ Third, regular and systematic eGFR measurements were not consistently obtained during the follow-up period after the initial assessment. Fourth, the proportion of catheter ablation as the initial rhythm control strategy in this study was only 0.5%, which was significantly lower than the reported 7% in the EAST-AFNET 4 trial.^3^ However, over the past decade, the number of ablation procedures has gradually increased.^32^ Consistent with this trend, among patients who underwent rhythm control strategies, 843 (5.4%) underwent the procedure within 1 year after the initiation of follow-up, and eventually, 1,473 (9.4%) underwent the procedure until the completion of follow-up in this study. Considering the supporting evidence of the superior effectiveness of catheter ablation compared to that of AAD in maintaining sinus rhythm and improving symptoms in patients with AF, further research is warranted to determine whether this analysis would yield different conclusions.^10, 11^

## Conclusions

In this nationwide population-based study, rhythm control strategies were associated with better cardiovascular outcomes than rate control strategies in patients with renal failure. In patients with ESRD, however, the beneficial association was less prominent, and the risk of composite safety outcome was higher than that of rate control strategies. Considering the increased incidence of side effects associated with rhythm control strategies, these findings will aid in the targeted selection of patients with AF and renal failure who may benefit from rhythm control strategies.

## Data Availability

Data for this retrospective analysis were obtained from the National Health Insurance Service (NHIS) database in Korea. The NHIS is managed by the Korean government and covers the majority (97.1%) of the Korean population, with the remaining 3% receiving medical aid. The database includes information on sociodemographic factors, inpatient and outpatient services, prescriptions, and mortality and is accessible to the public on the NHIS website (http://nhiss.nhis.or.kr).

## Acknowledgements

We thank the National Health Insurance Service of Korea for their cooperation.

